# Language deficits across PET-based Braak stages of tau accumulation in Alzheimer’s disease

**DOI:** 10.1101/2025.09.20.25336243

**Authors:** Anna Marier, Jaime Fernández Arias, Étienne Aumont, Brandon J. Hall, Arthur C. Macedo, Nesrine Rahmouni, Gleb Bezgin, Paolo Vitali, Pedro Rosa-Neto, Maxime Montembeault

## Abstract

**INTRODUCTION:** Widespread language complaints in the cognitively unimpaired (CU) may reflect Alzheimer’s Disease (AD) pathology and future objective impairments.

**METHODS:** 138 CU, 45 mild cognitive impairment and 28 dementia participants from the TRIAD cohort underwent ^18^F-MK-6240 tau-PET and ^18^F-AZD-4694 amyloid-PET. Word-finding complaints, confrontation naming, semantic and phonemic fluency and word-knowledge were evaluated. Covariance, direct and stepwise discriminant, and voxel-wise regression analyses were conducted.

**RESULTS:** Word-finding complaints appeared in early tau stages (Braak 1–2), followed by naming difficulties (Braak 3–4), and widespread language impairments in later stages (Braak 5–6). Complaints over forgetting the names of objects, naming, and APOE significantly improved classification of early AD pathology. In CU, complaints over forgetting names of objects were linked to left fusiform and inferior temporal gyri tau accumulation.

**DISCUSSION:** Language measures are useful in detecting and tracking AD-related pathophysiologies. Results encourage refinement of clinical tools for early detection and disease monitoring.

**Highlights:** Language decline parallels tau buildup across PET-based Braak stages of AD.

Subjective anomia marks earliest tau-related language symptom (Braak 1–2).

Objective naming deficits emerge in the middle tau spread stages (Braak 3–4).

Advanced tau spread reflects significant and widespread language impairments.

Word-finding complaints correlate with left fusiform and inferior temporal tau.

**Research in context:** **Systematic review:** The literature was reviewed using traditional sources. The core biological definition of Alzheimer’s disease (AD) has recently been linked to its defining cognitive clinical features of episodic memory impairments. Widespread subjective language complaints amongst cognitively unimpaired (CU) older adults and objective language impairments observed across the AD continuum suggests these measures and the further bridging of biological and clinical definitions of AD could play a critical, cost-effective role in disease detection and monitoring.

**Interpretation**: Results extend to tau previous findings describing language changes in AD and related to amyloid status and grey-matter atrophy. They also establish the likely staging of language impairments across the biological AD continuum.

**Future directions:** The manuscript contextualises the use of subjective word-finding complaints, alongside genetic risks to significantly enhance the prediction of underlying AD related pathology in CU. Languages measures used in clinical practice remain limited however and better test should be utilized and developed.

## 1. Background

Alzheimer’s disease (AD) has long been defined clinically, with core features including episodic memory impairment, language and visuospatial deficits, executive dysfunction, behavioral or personality changes, as well as a clear impact on daily functioning (McKhann et al., 2011). A more recent definition instead bases itself primarily on the disease’s underlying biological processes. At their core, amyloid beta (Aβ) and tau proteins accumulation within the brain is understood to be a leading cause of neuronal dysfunction and degeneration (Jack et al., 2024). Under this new framework, disease progression can be understood as a hierarchical progression of accumulating tau throughout brains identified with pathological levels of Aβ (Braak et al., 2006; Macedo et al., 2023). It is this progression which ultimately leads to cognitive symptoms (Jack et al., 2024). Recent efforts have contributed to the integration of episodic memory, as it was understood under clinically defined AD, within the framework of its biological underpinnings (Fernández Arias et al., 2023). Other core AD cognitive domains have, however, yet to be similarly integrated to the biological framework, in spite of clinical diagnosis of dementia requiring impairments in at least two (McKhann et al., 2011).

Previous work, which has primarily focused on clinically defined AD, has highlighted potential links between language measurements and AD continuum progression. Naming abilities have for example been found to be impaired from the mild cognitive impairment (MCI) stage of AD (Joubert et al., 2021) and were related to grey matter (GM) atrophy within the left anterior temporal lobe (Frings et al., 2011; Domoto-Reilly et al., 2012) and left inferior prefrontal gyrus (Joubert et al., 2010). Verbal fluency has likewise seen groups of patients impaired from the MCI stage (Nutter-Upham et al., 2008; Chasles et al., 2020) or further along the AD continuum (particularly letter fluency; Weakley et al., 2014) and has been related to GM atrophy within the left inferior temporal gyrus, left insular cortex, bilateral hippocampus and parahippocampal gyrus, extending also, in the case of semantic fluency, to the anterior and posterior cingulate cortices, as well as bilateral caudate and cerebellar crus II regions (Rodríguez-Aranda et al., 2016). Impairments on word-knowledge tasks centered around accuracy of definitions were also found in dementia due to AD (Gontkovsky et al., 2022). When moving earlier in the continuum, subjective word-finding complaints were found to be amongst the most commonly reported in cognitively unimpaired older adults (Montembeault et al., 2022), with such complaints being linked to atrophy in the left fusiform gyrus and linked to elevated levels of Aβ (Jessen et al., 2014; Montembeault et al., 2022).

Despite clear links between language impairments and AD-related GM atrophy, little is known of deficits’ progression across the tau-based biologically defined AD continuum, nor of vulnerable language-related locations for pathological accumulation of tau proteins within the brain. This subject is particularly challenging considering the wide range of protective and risk factors involved (Alzheimer’s disease facts and figures, 2025) some of which are more commonly controlled for, such as age, sex, education and genetics, although even then, their impacts are too often excluded from interpretation of results. Another important challenge is that cost-effective and early detection is paramount, not only because of the costly and insidious nature of the disease, but also because 68% of its projected increase by 2050 is expected to burden low- and middle-income countries (Alzheimer’s disease facts and figures, 2025).

This article therefore aims to (a) assess whether language performance measures can significantly differentiate participants across the tau progression continuum of biologically defined AD using individual language tests; or (b) a combination of language and demographic measures; (c) investigate upon the significant relationships between language performances and location of pathological protein accumulations within the brain; and (d) inform on the most useful language and demographic measures for the classification of early AD pathology. We hypothesize that (1) participants at the earliest stages of pathology on the biologically defined AD continuum will present elevated word-finding complaints, followed by impaired confrontation naming and semantic fluency performances in the middle stages, and phonemic fluency and verbal comprehension impairments in the later stages; (2) important demographic measures such as age, sex, education and genotype will modulate our understanding of group differences; (3) pathological protein accumulation associated with language impairment will primarily be centered around the left temporal lobe and other previously highlighted brain regions; and (4) word-finding complaints and important predictor measures, such as genotype, will be most useful in the classification of early AD pathology.

## 2. Methods

The data used in the preparation of this article were obtained from the Translational Biomarkers of Aging and Dementia (TRIAD; Stevenson et al., 2020; https://triad.tnl-mcgill.com/cohort-description/) and, to a much lesser extent, the Head-to-head Evaluation of tau tracers in Alzheimer’s Disease (HEAD; Lussier et al., 2024; https://head-study.info/head-cohort) cohorts databases.

### 2.1. Participants

Whilst Braak staging and amyloid-β positivity formed the primary, biology-focused lens through which this study was conducted, clinical presentation was used to better describe the sample and to orient the interpretation of results. Clinical diagnosis was performed before biomarker data was collected and was established through consensus reached by a multidisciplinary team composed of neuropsychologists, physicians and nurses.

Participants were classified as cognitively unimpaired (CU) if they showed no objective cognitive impairment and a Clinical Dementia Rating (CDR) total score of 0. Participants classified with mild cognitive impairment (MCI) presented subjective and objective cognitive impairments in at least one of the recognized AD continuum domains (that is episodic memory, language, visuospatial, executive function or behavioral abnormalities or personality changes; McKhann et al., 2011) and a CDR total score of 0.5. Their general cognition, functional performance and independence in activities of daily living were sufficiently preserved so that a diagnosis of AD could not be made.

Participants classified with Dementia due to Alzheimer’s Disease (AD) presented more significant cognitive impairments in at least two of the recognized AD continuum domains which in this case interfered with independence and activities of daily living, and a CDR total score equal or superior to 0.5 (McKhann et al., 2011).

Cohorts exclusion criteria included (a) inability to speak French or English; (b) sensory impairment hampering neuropsychological evaluation; (c) recent major surgery or traumatic brain injury; (d) magnetic resonance imaging (MRI) or positron emission tomography (PET) contraindications; (e) neurological, psychiatric or systemic conditions not adequately controlled through a stable medication regimen; (f) ongoing substance abuse; and (g) unavailability of reliable informant, such as a family member or close friend (Pascoal et al., 2021; Macedo et al., 2024^a^).

In addition to cohorts’ exclusion criteria, we applied for this study 9 additional criteria. Starting with 1244 longitudinal time points, we excluded those which (h) did not present language performance data, retaining cases presenting limited data (499 removed); (i) did not have ^18^F-MK-6240 tau-PET scan conducted or had scans conducted that did not pass quality control (119 removed); (j) stood outside the Alzheimer continuum diagnostic groups of interests for this study (those classified as CU young, MCI not due to AD, atypical AD, suspected Parkinson’s or those with suspected non-Alzheimer pathophysiology; 178 removed); (k) did not have Braak staging determination conducted (30 removed); (l) at Braak stage 0, did not have Aβ status confirmed (57 removed) or at Braak stages 1 to 6, had negative Aβ status (60 removed); (m) stood outside Montréal Cognitive Assessment (MoCA)-based, sex and education adjusted CU normative threshold for cognitive impairment (for the Quebec population; below -1.31 z-score; Larouche et al., 2016), or CU with missing MoCA (6 removed); (n) had multiple time points left after the previous step, we prioritized retaining earliest time points for this selection (82 removed); and finally (o) departed significantly from their Braak stage groups in terms of language performance, evidenced by what turned out to be a combination of within-group extreme univariate (single test performance deviating beyond 3.29 Braak subgroup z-score; Tabachnick and Fidell, 2019) and multivariate abnormality (Mahalanobis distance based on our 6 language variables of interest with *p* < .001 alpha threshold: critical χ2 = 22.45; Tabachnick and Fidell, 2019; Kline, 2023), which we believe to be indicative of language impairment unrelated to AD pathology (two Braak 0 CU removed).

After all considerations, 211 uniquely time pointed participants remained, of which 138 CU, 45 with MCI due to AD, and 28 with dementia due to AD.

### 2.2 Procedure

#### 2.2.1 Cognitive assessments

##### Everyday Cognition Questionnaire

Subjective cognitive complaints regarding one’s word-finding abilities were assessed using the self-reported Everyday Cognition questionnaire (ECog; Farias et al., 2008). This 39-item tool probes participants for perceived changes in their cognitive abilities across domains of memory (eight items), language (nine items), visuospatial/perceptual (seven items); and executive function abilities of planning (five items), organization (six items) and divided attention (four items). Participants report that, relative to 10 years prior, a given ability has evolved to be (a) better or not changed; (b) questionable or occasionally worse; (c) consistently a little worse; (d) consistently much worse; or finally that they (e) do not know. Based on a previous study, we selected the most frequently reported language complaints in CU older adults, that is ECog 1: “forgetting the names of objects” and ECog 3: “Finding the right words to use in a conversation” (Montembeault et al., 2022).

##### Boston Naming Test

Objective confrontation naming was assessed using the Boston Naming Test (BNT; Kaplan et al., 2001). This originally 60-item test measures one’s ability to orally label (name) drawings of objects. In order of frequency, from most to least commonly known, participants are presented with a black and white image and have 20 seconds to name what the drawing represents. Six consecutive failures lead to the discontinuation of the test. A semantic cue is given if the participant fails to recognize the picture (e.g., answering bench instead of tree) or if they state that they do not know what the picture represents. The semantic cue is either a short explanation about the item (e.g., for a mask: “it’s part of a carnival fantasy”) or a superordinate category (e.g., for a beaver: “it’s a kind of animal”). One score is recorded before and another after the cue is presented. For the purpose of this study, we used the former, “raw” score. However, the cue remained useful for participants’ continuation of the test. The present cohorts only administered odd numbered items on the standard 60 item BNT, which gives a maximum score of 30.

##### Delis-Kaplan Executive Function System: letter and category fluency subtests

Phonemic and semantic verbal fluency were assessed using the Delis-Kaplan Executive Function System (Delis et al., 2001). One subtest of this larger battery is a letter (phonemic) fluency test during which participants are given 60 seconds to generate as many words as they can, beginning with a designated letter. During a test session, participants will go through 3 different letters (60 sec. × 3), that is “P, F, L” for French and “F, A, S” for English. Repetitions are not counted towards the total words produced. We used the sum of words correctly produced over all trials as our measure of phonemic fluency performance.

Another subtest of this battery is a category (semantic) fluency test during which participants are once again given 60 seconds to generate as many words as they can, belonging, this time, to a designated semantic category. During a test session, participants will go through 2 different semantic categories, that is “animals’ names” and “boys’ names” (60 sec. × 2). Repetitions are again not counted. We used the sum of words correctly produced over both trials as our measure of semantic fluency performance.

##### Wechsler Abbreviated Scale of Intelligence-II: vocabulary subtest

Word-knowledge was assessed using the Wechsler Abbreviated Scale of Intelligence’s second edition (WASI-II; Wechsler, 2011). One subtest of this larger scale is a 31-item “vocabulary” subtest conducted in two parts. First, a 3-item picture confrontation naming test, in which participants are asked to name visually presented objects, receiving either “correct” (1) or “incorrect” (0) scores based on their answer. Second, a 28-item word-knowledge test in which participants are asked to define orally and visually presented words, receiving a score in accordance with the accuracy and quality of their definition (either 0, 1 or 2), following the criteria outlined in the test manual, with some answers prompting queries from the examiner for additional details. Three consecutive failures lead to the discontinuation of the test. We used the sum of all items as our measure of word-knowledge.

#### 2.2.2 Neuroimaging

##### Structural magnetic resonance imaging acquisition and processing

To obtain anatomical images of the brain, structural MRI was conducted using a 3T Siemens MAGNETOM scanner equipped with a standard head coil. A Magnetization Prepared Rapid Gradient Echo sequence was employed (repetition time of 2300 milliseconds, echo time of 2.96 ms, 9°flip angle, coronal orientation perpendicular to the double spin echo sequence, 1 × 1 mm^2^ in-plane resolution of 1 mm slab thickness; Ferrari-Souza et al., 2022, see supplementary) to obtain brain wide 1mm isotropic voxels with T1-weighted contrast which were then corrected for field distortions and non-uniformity, followed by parcellation into substructures (using FreeSurfer; https://surfer.nmr.mgh.harvard.edu/; Fischl et al., 2004), with regions of interest defined according to the Desikan—Killiany—Tourville (DKT) atlas (Desikan et al., 2006), and segmentation into probabilistic types of tissue maps (using Statistical Parametric Mapping 12 segmentation tool; https://fil.ion.ucl.ac.uk/spm/; Ashburner et al., 2003), using a Montréal Neurological Institute

(MNI) Translational Neuroimaging Laboratory (TNL) in-house pipeline. GM probability maps were then non-linearly transformed to the Alzheimer’s Disease Neuroimaging Initiative (ADNI) reference space (using Advanced Normalization Tools; ANTs; https://stnava.github.io/ANTs/; Avants et al., 2011). Resulting images were then finally smoothed using an 8mm full-width half-maximum Gaussian kernel, followed by visual inspection to ensure proper alignment to the ADNI template (Ferrari-Souza et al., 2022, see supplementary).

##### Amyloid-PET and Tau-PET acquisition and processing

To locate and measure AD-related pathological protein aggregation in the brain, PET scans were conducted using a brain-dedicated Siemens High Resolution Research Tomograph.

^18^F-MK-6240 radioligand was used to image tau neurofibrillary tangles aggregation, with acquisition proceeding 90 to 110 minutes following intravenous bolus injection. Scans were reconstructed using an ordered-subsets expectation maximization (OSEM; Hudson and Larkin, 1994) algorithm on a four-dimensional volume with four frames (4 × 300 seconds; Pascoal et al., 2018; Pascoal et al., 2020).

^18^F-AZD-4694 radioligand was used to image amyloid-β plaques aggregation, with acquisition proceeding 40 to 70 minutes following intravenous bolus injection. OSEM was once again used, this time on a four-dimensional volume with three frames (3 × 600 seconds; Cselényi et al., 2012; Therriault et al., 2021).

For attenuation correction, PET acquisition was followed by a 6-min transmission scan conducted with a rotating ^137^Cs point source. Images were additionally corrected for motion, dead time, radioactive decay as well as random and scattered coincidences and were also meninges and skull stripped (Pascoal et al., 2020). PET registration to corresponding T1-weighted MRI images was finalized, once again using a MNI TNL in-house pipeline, by linear registration of PET images to native T1 space (using ANTs; Avants et al., 2011) and non-linear transformation to ADNI reference space (using the transform obtained from the MRI processing step). The resulting PET images were likewise smoothed using the same 8mm full-width half-maximum Gaussian kernel.

To quantify radioligand binding, standardized uptake value ratios (SUVR; Lucignani et al., 2004) were computed using the (a) inferior cerebellar grey matter as a reference region in the case of tau-PET ^18^F-MK-6240 (Pascoal et al., 2020); and (b) whole cerebellar grey matter in the case of amyloid-PET ^18^F-AZD-4694 (Cselényi et al., 2012).

##### Amyloid positivity and PET-Braak staging

Individuals were categorized based on previously established pathological thresholds of protein accumulation.

Amyloid-β pathology was classified using a dichotomous threshold (negative [–]/positive [+]), with individuals considered “amyloid-β positive” (Aβ+) when ^18^F-AZD-4694 SUVR composite average across the precuneus, prefrontal, orbitofrontal, parietal, temporal, and cingulate cortices exceeded 1.55 (Therriault et al., 2021).

Tau pathology was classified using a tau-based Braak staging system for AD (Braak et al., 2006; Macedo et al., 2023). This system is based on Braak’s proposed framework, which describes typical AD-related tau aggregation in its hierarchical movement throughout the brain following disease progression, with each stage characterized by specific and accumulating regional abnormalities. In TRIAD, this was defined as beginning in the transentorhinal cortex (Braak stage 1); and progressing thereafter onto the entorhinal cortex and hippocampus (Braak stage 2); the amygdala and parahippocampal, fusiform, and lingual gyri (Braak stage 3); the insular cortex, lateral temporal lobe, posterior cingulate cortex, and inferior parietal lobule (Braak stage 4); the orbitofrontal cortex, superior temporal and inferior frontal gyri, cuneus, anterior cingulate cortex, supramarginal gyrus, lateral occipital cortex, precuneus, superior parietal lobule, superior frontal gyrus, and rostromedial frontal cortex (Braak stage 5); ultimately extending to the paracentral lobule, postcentral and precentral gyri and pericalcarine cortex (Braak stage 6; Pascoal et al., 2020; Therriault et al., 2022) and beyond. An absence of such abnormalities was categorized as “Braak 0”. Upon reaching any Braak stage and to be considered representative of the “typical progression” of tau accumulation, one must have surpassed abnormality thresholds in previous Braak stages. Abnormality thresholds were defined as 2.5 standard deviation above mean SUVR of CU individuals younger than 26 years old (Pascoal et al., 2020; Pascoal et al., 2021).

With regards to this study, participants were categorized based on their Aβ status, with segregated Aβ— and Aβ+ individuals of Braak stage 0, and, like previously mentioned, similar to other works before us, rejected Aβ— individuals of Braak 1 through 6 (Jack et al., 2018; Macedo et al., 2024^b^). Similarly again, to increase the sample size of each Braak-amyloid subgroup, we used a simplified Braak stage grouping, resulting in the following final subgroups: Braak 0 Aβ– (*N* = 100), Braak 0 Aβ+ (*N* = 25), Braak 1–2 Aβ+ (*N* = 35 [9/26]), Braak 3–4 Aβ+ (*N* = 24 [6/18]) and finally, Braak 5–6 Aβ+ (*N* = 27 [10/17]).

### 2.3 Statistical analyses

#### 2.3.1 Behavioral analyses

To describe the sample, Fisher’s exact test was used to assess between-subgroup pairwise categorical differences, that is, of sex, primary language spoken, handedness and APOE genotype, applying Bonferroni correction to adjust for multiple comparisons. This was done using dichotomous presence of APOE ε4 (yes/no) to prevent issues that arise when categories have too few observations (Howell, 2007). However, in subsequent analyses, APOE genotype was computed ordinally, as follows, to reflect the established protective impact of carrying ε2 alleles as well as the cumulative risk of carrying ε4 alleles on the likelihood of developing late-onset AD: (one) ε2 | ε3 with least risks; followed by (two) ε3 | ε3; (three) ε3 | ε4; and finally (four) ε4 | ε4 with most risks (Qian et al., 2017; Reiman et al., 2020; Sienski et al., 2021). One-way analyses of variance (ANOVA) and Tukey post-hoc tests were used for all other (continuous) variables, that is age, education, CDR, MMSE, MoCA and amyloid-PET ^18^F-AZD-4694 full cerebellum GM SUVR composite.

To test subgroup differences in terms of language performance, we modeled a number of multiple linear regressions that predicted a given language score based on Braak subgroups, extracting an ANOVA table (with Tukey post-hoc) to test for the factor as a whole, and controlling for (a) sex; (b) age; (c) education; (d) primary language spoken (French or English); (e) handedness (left or right) and (f) ordinally-computed APOE genotype. Epsilon Square was used as a measure of effect size (ε^2^; Okada, 2013). This can also be understood as a series of analyses of covariance (ANCOVA).

To test the number of dimensions along which Braak subgroups differed significantly, subgroup location along discriminant functions, predictor relationships to discriminant functions and to enable us to understand how dimensions of differences can be interpreted and also to test for variance explained by multivariate modeling of all of our variables, we conducted a direct discriminant analysis using the “candisc” R package (Friendly and Fox, 2024). Overall Pillai’s trace was extracted from the multivariate linear model using the “Anova” function provided by the “car” R package (Fox and Weisberg, 2019). Overall variance explained was calculated as canonical variables *R*^2^ sum, divided by canonical variable sum (Cramer and Nicewander, 1979). Our model included (a) age; (b) sex; (c) education; (d) primary language spoken; (e) handedness; (f) APOE genotype; (g) Subjective word-finding complaints evaluated with the Ecog language question 1: Forgetting the names of objects; (h) Subjective word-finding complaints evaluated with the Ecog language question 3: Finding the right words to use in a conversation; (i) BNT total score; (j) WASI-II: Vocabulary total score; (k) semantic fluency; and (l) phonemic fluency.

To evaluate predictor variable contribution regarding early AD pathology (Aβ+ and tau), we conducted a forward stepwise discriminant analysis using the “folda” R package (Wang and Yao, 2025) with Pillai’s trace set as test for forward selection and alpha setting of .05. Variables were the same as with the previous model. However, we additionally included MoCA total score as a predictor variable for comparison of usefulness in the context of early AD pathology classification and screening.

All behavioral statistical analyses were performed using R Statistical Software (version 4.4.1 for Windows operating system; R Core Team, 2024).

#### 2.3.2 Neuroimaging analyses

To evaluate the associations between language variables and in vivo aggregation of tau proteins in the brain, voxel-wise regression analyses were conducted using the “RMINC” R package (version 1.5.2.3; Lerch et al., 2017). Our linear models predicted voxel-wise tau-PET SUVR using a given language score and controlling for (a) age; (b) sex; (c) education; (d) primary language spoken; (e) handedness; (f) APOE genotype; and (g) amyloid-PET ^18^F-AZD-4694 full cerebellum GM SUVR composite and corrected for multiple comparisons using a random field theory (Brett et al., 2004) cluster threshold of *p* < .001. To better interpret regions of tau aggregation, we checked for overlap between the significant T-statistical parametric maps and lateralized DKT atlas reference substructures.

All neuroimaging analyses were performed using R Statistical Software (version 3.6.1 for Ubuntu, a Linux-based operating system; R Core Team, 2024).

## 3. Results

Demographic characteristics of our sample are presented in table 1. Tau-PET-based Braak stages of AD subgroups differed with regards to age (*F* [4, 206] = 2.39, *p* = .05, ε^2^ = 0.03) and APOE genotype (based on the presence of at least one ε4 allele; χ^2^ [4] = 48.81, *p* < .001). Descriptive clinical characteristics are also presented in table 1. Groups differed with regards to disease severity as evaluated by the CDR (*F* [4, 197] = 54.15, *p* < .001, ε^2^ = 0.51), global cognition as measured by the MMSE (*F* [4, 197] = 30.82, *p* < .001, ε^2^ = 0.37) and MoCA (*F* [4, 197] = 61.06, *p* < .001, ε^2^ = 0.54) as well as Amyloid-PET load (based on^18^F-AZD-4694 full cerebellum GM SUVR composite; *F* [4, 206] = 124.77, *p* < .001, ε^2^ = 0.70), all of which followed expected AD continuum progression.

**Table 1:**
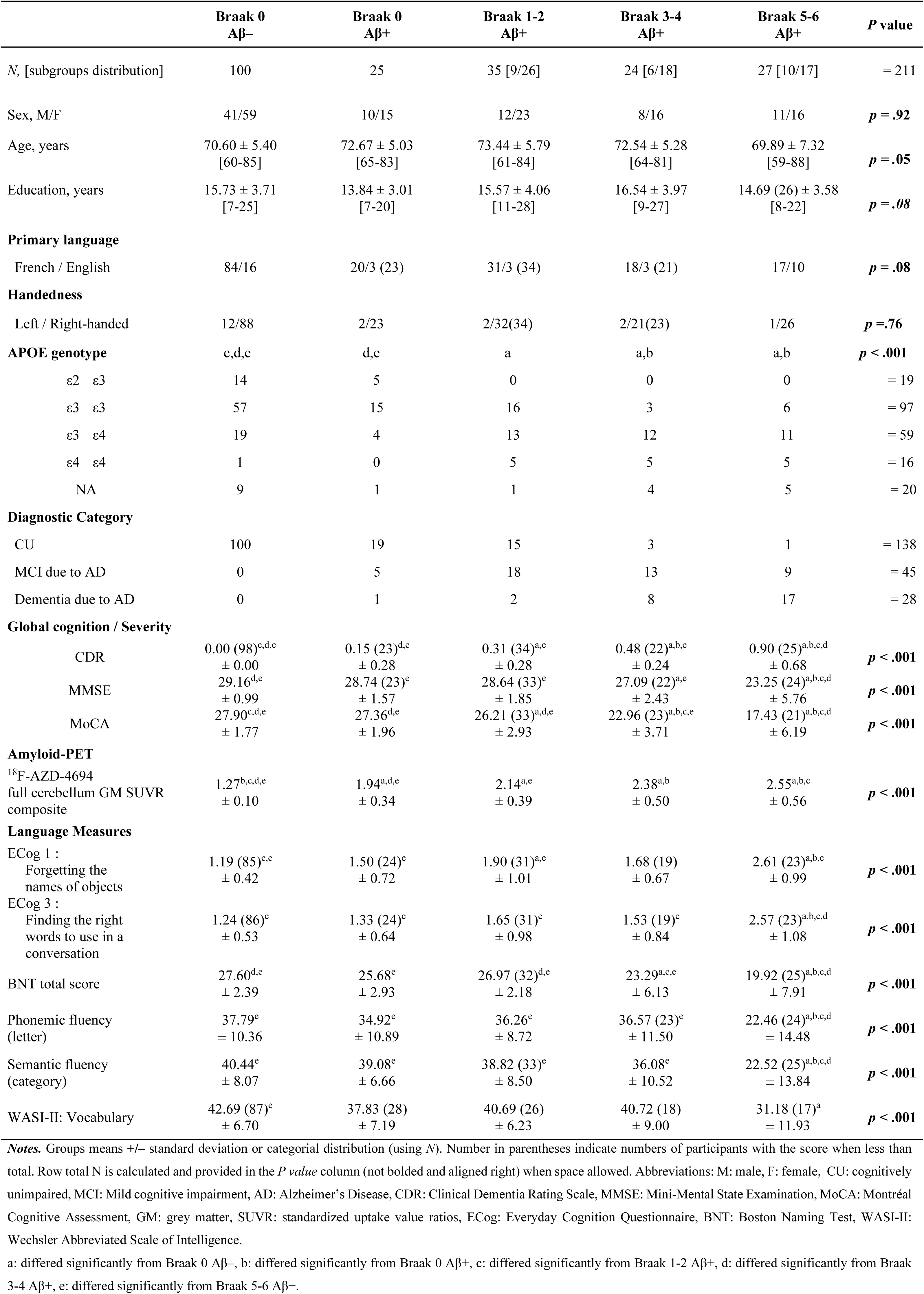
Demographic, clinical and language characteristics across subgroups

### 3.1 Language impairment across Tau-PET-based Braak stages of AD

As presented in table 1 and figure 1, when controlling for sex, age, education, primary language spoken, handedness and ordinally-computed APOE genotype, groups significantly differed according to their levels of subjective word-finding complaints over forgetting the names of objects (ECog 1; *F* [4, 149] = 15.95, *p* < .001, ε^2^ = 0.28) and over finding the right words to use in a conversation (ECog 3; *F* [4, 150] = 13.25, *p* < .001, ε^2^ = 0.24) as well as according to their BNT (*F* [4, 168] = 21.63, *p* < .001, ε ^2^ = 0.32), phonemic fluency (*F* [4, 170] = 9.17, *p* < .001, ε ^2^ = 0.16), semantic fluency (*F* [4, 169] = 18.18, *p* < .001, ε ^2^ = 0.28) and WASI-II: vocabulary (*F* [4, 135] = 8.21, *p* < .001, ε ^2^ = 0.32) performances. In some cases, a slight departure from homogeneity of variance between groups suggested a more conservative approach could be employed in the interpretation of results. Considering this, we followed Tabachnick and Fidell (2019)’s recommendation of halving the *p* value significance thresholds from .05 to .025, .01 to .005 and the .001 to .0005. However, we found that this did not impact any of the obtained ANCOVA results. Overall, comparatively to the biomarker-negative individuals (Braak 0 Aβ–), those who had reached Braak 1-2 Aβ+ presented significantly elevated word-finding complaints over forgetting the names of objects (ECog 1), whilst those who had reached Braak 3-4 Aβ+ presented impairments in confrontation naming (BNT) and those who had reached Braak 5-6 Aβ+ presented elevated subjective word-finding complaints (ECog 1 and 3) alongside impairments in all objective language measures (confrontation naming, fluencies, word-knowledge). Comparatively to amyloid-positive individuals with no pathological tau progression (Braak 0 Aβ+), those who had reached Braak 5-6 Aβ+ presented relatively preserved word-knowledge (WASI-II) and otherwise presented elevated subjective word-finding complaints alongside impairments in all other objective language measures (naming, fluencies).

**Figure 1:**
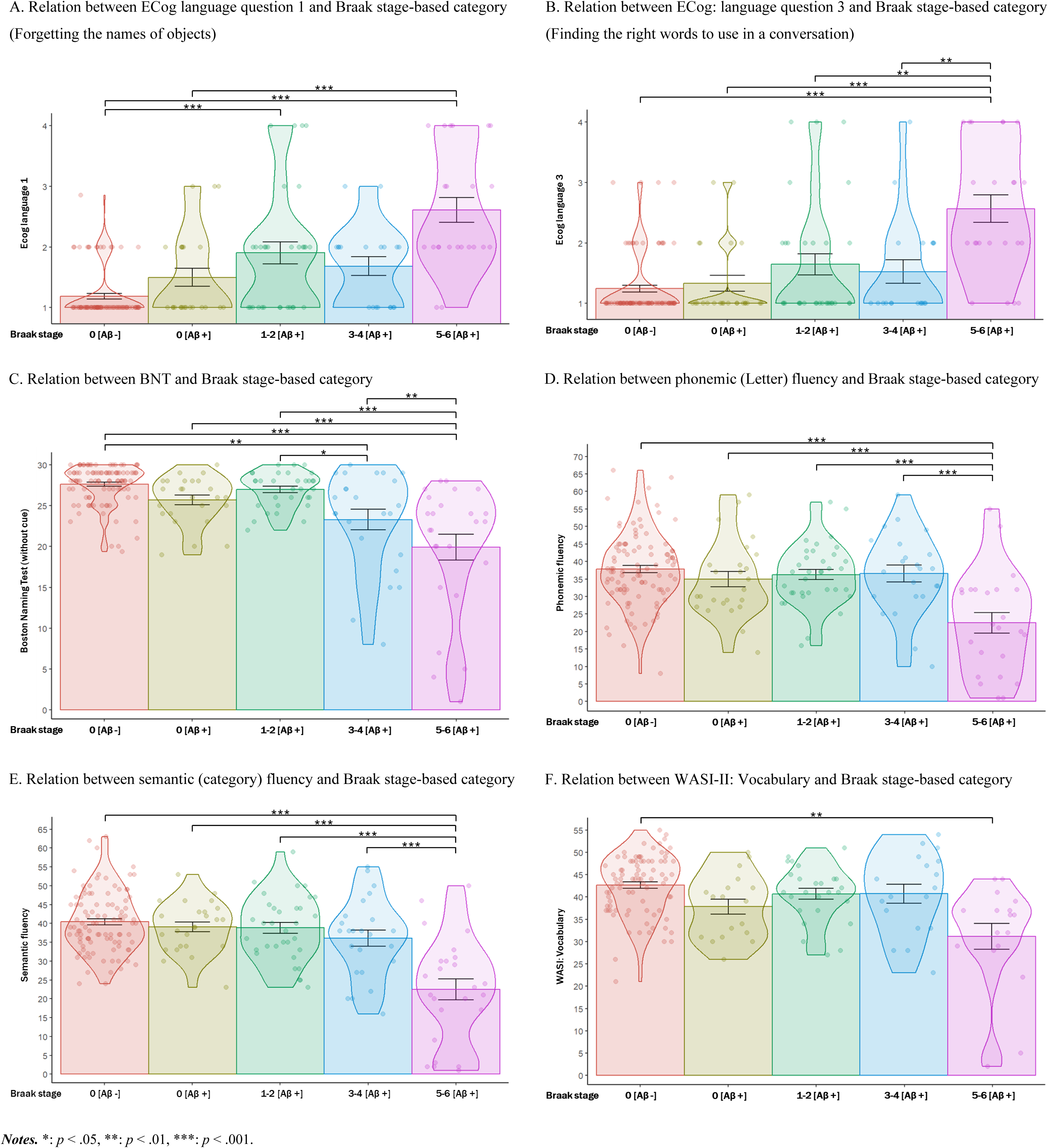
Language deficits across the biologically defined AD continuum

Comparatively to individuals in Braak 1-2 Aβ+, those who had reached Braak 3-4 Aβ+ presented impairments in confrontation naming and those who had reached Braak 5-6 Aβ+ presented elevated word-finding complaints over finding the right words to use in a conversation alongside objective impairments in confrontation naming as well as verbal fluencies. Finally, comparatively to individuals in Braak 3-4 Aβ+, those who had reached Braak 5-6 Aβ+ presented elevated word-finding complaints over finding the right words to use in a conversation alongside objective impairments in confrontation naming as well as verbal fluencies.

A second dimension revolved primarily around a negative and strong relationship to Braak 0 Aβ+, with, in order of their importance, relatively weak and positive associations to Braak 0 Aβ–, weaker and negative associations to Braak 1-2 Aβ+ and positive associations to Braak 5-6 Aβ+. Notable associated variables were, in order of their importance, age alongside confrontation naming, WASI-II: Vocabulary total score, education, complaints over forgetting the names of objects and handedness. Following this second dimension’s subgroup associations, we gave it the interpretative name of “Braak 0 Aβ+ profile”.

A third significant dimension of group differences (although least explicative of total variance) revolved primarily around a dual relationship between a negatively associated Braak 3-4 Aβ+ and a positively associated Braak 5-6 Aβ+. Notable associated variables were, in order of their importance, education alongside WASI-II: Vocabulary total score, elevated, complaints over finding the right words to use in a conversation, semantic and phonemic fluencies as well as complaints over forgetting the names of objects. Following this third dimension’s subgroup associations, we gave it the interpretative name of “Braak 3-4 vs Braak 5-6”.

Next are multivariate analyses results, with table 2 and supplementary material figure 1 presenting dimensions along which group differences could be interpreted. Unlike with the previous set of results, all language variables were simultaneously evaluated in a model inclusive of the interpretable impact of covariates of sex, age, education, primary language spoken, handedness and ordinally-computed APOE genotype.

**Table 2:** Dimensions along which the biologically defined AD continuum can be understood.

| Canonical variable | 1 | 2 | 3 |
| --- | --- | --- | --- |
| Interpretative name | “Proportional to tau spread” | “Braak 0 Aβ+ profile” | “Braak 3-4 vs Braak 5-6” |
| Overall Pillai’s trace = .98, $F_{approx}(48, 452) = 3.08, p < .001$ | | | |
| Wilks’ Λ | .30 | .57 | .74 |
| $F_{approx}$ | (48, 426)<br>= 3.28 | (33, 328)<br>= 2.07 | (20, 224)<br>= 1.78 |
| P value | < .001 | < .001 | < .01 |
| $(R^2 \text{ sum} / 3) \approx$<br>29% variance explained across all | .48 | .23 | .17 |
| Braak 0 [Aβ negative] | 0.64 | 0.35 | 0.01 |
| Braak 0 [Aβ positive] | 0.58 | -1.28 | 0.11 |
| Braak 1-2 [Aβ positive] | -0.46 | -0.23 | 0.04 |
| Braak 3-4 [Aβ positive] | -1.43 | -0.01 | -1.21 |
| Braak 5-6 [Aβ positive] | -2.10 | 0.21 | 0.81 |
| ❖ Advanced age | 0.05 | -0.39 | -0.15 |
| ❖ Higher proportion of men | 0.11 | -0.20 | 0.22 |
| ❖ Advanced education | -0.06 | 0.32 | -0.45 |
| ❖ Higher proportion of English speakers | -0.10 | 0.08 | 0.06 |
| ❖ Higher proportion of left-handed | 0.16 | 0.27 | -0.01 |
| ❖ Higher APOE genotype associated risk | -0.76 | 0.17 | -0.29 |
| ❖ Elevated complaints, ECog 1:<br>Forgetting the names of objects. | -0.69 | -0.30 | 0.39 |
| ❖ Elevated complaints, ECog 3:<br>Finding the right words to use in a conversation. | -0.59 | 0.04 | 0.44 |
| ❖ Better BNT total score | 0.51 | 0.39 | 0.17 |
| ❖ Better phonemic fluency | 0.23 | 0.04 | -0.41 |
| ❖ Better semantic fluency | 0.40 | -0.08 | -0.42 |
| ❖ Better WASI-II: Vocabulary total score | 0.29 | 0.36 | -0.45 |

**Notes.** This multivariate model of language, demographic and genetic measures for the classification of AD pathology subgroups along the tau accumulation continuum was built using results from a discriminant analysis. Coloring was applied to cells containing values according to the direction of the relationship (red for negative, green for positive with yellow in between for unrelated relationship) and the relative strength of other cells. This was done in two steps (1) for the relationships of the biological stages with the Canonical variables; and (2) for the relationships of the language, demographic and genetic measures with the Canonical variables. Colors in “1” are therefore not dependent on the same scaling as colors in “2”. Additional note for the uninitiated: be mindful of double negatives, which can sometimes be difficult to interpret.

The most important dimension of group differences followed the pattern of tau accumulation within our sample. In other words, Braak 0 Aβ– and Aβ+ were similarly and positively related to this first dimension, followed by a negative and mild relationship to Braak 1-2 Aβ+, a negative and much stronger relationship to Braak 3-4 Aβ+ and negative and even stronger relationship to Braak 5-6 Aβ+. Notable associated variables were, in order of their importance, APOE-associated risk of developing AD, complaints over forgetting the names of objects, complaints over finding the right words to use in a conversation, confrontation naming, and semantic fluency. Following this first dimension’s subgroup associations, we gave it the interpretative name of “proportional to tau spread”.

Additional note for the uninitiated: be mindful of double negatives, which can sometimes be difficult to interpret.

### 3.2 Language impairment association to tau aggregation within the brain

Whole sample-based masks of significant associations between language scores and tau-PET are presented in figure 2, with models’ statistics and overlapping with lateralized DKT atlas reference substructures masks presented in table 3. Results presented control for age, sex, education, ordinally-computed APOE genotype, primary language, handedness and amyloid-PET ^18^F-AZD-4694 full cerebellum GM SUVR composite. Substructure mapping reference can be found in supplementary material figure 2. Overall, language measures were associated with large and diffuse positive clusters of tau aggregation across AD continuum brains. Relative to other measures, however, word-finding complaints relationships were by far most concentrated in the left hemisphere, with clear left frontal temporal T-statistical peaks (in both cases, even more so for ECog 1, which presented the least diffuse relationships to tau). Naming relationships extended otherwise further to bilateral frontal parietal regions, with clear left orbitofrontal and left superior frontal peaks. Regarding fluency measures, relationships to tau presented clear left posterior cingulate and left middle frontal peaks, with an otherwise relative balance between left and right hemispheres. Semantic fluency more particularly, additionally presented left superior temporal peaks. WASI-II: Vocabulary presented the second least diffuse clusters behind ECog 1 and in this case relationships to tau were largely centered around occipital, temporal and frontal regions with peaks in the left isthmus cingulate and bilateral rostral middle frontal.

**Figure 2:**
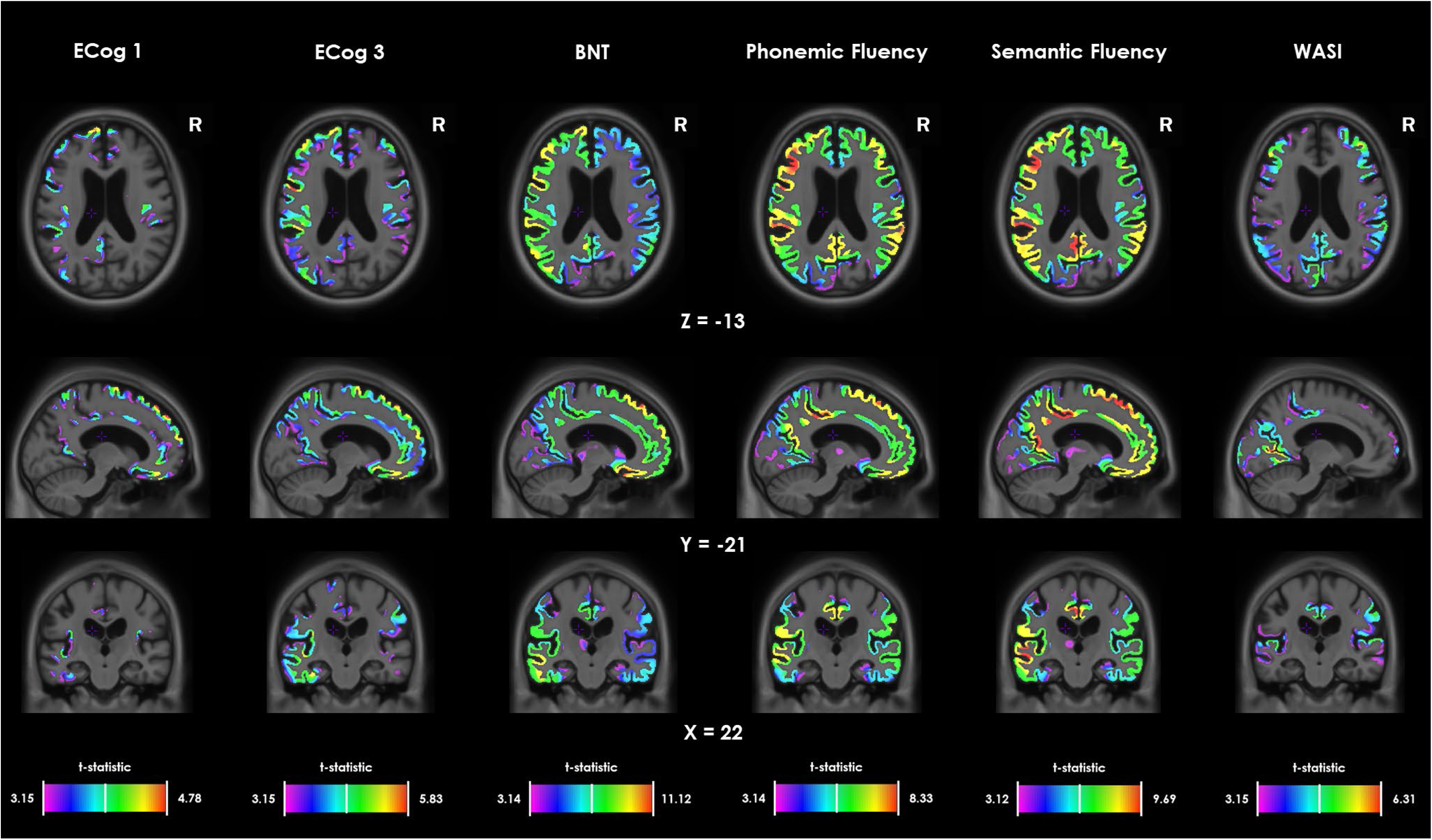
Whole sample – Location of significant associations between language scores and tau-PET

**Table 3:** Whole sample – Regions of significant associations between language scores and tau-PET

|  | ECog language 1 |  | ECog language 3 |  | Boston Naming Test |  | Phonemic Fluency |  | Semantic Fluency |  | WASI-II Vocabulary |  | Maximal possible<br>DKT Atlas Overlap<br>in number of voxels |  |
| --- | --- | --- | --- | --- | --- | --- | --- | --- | --- | --- | --- | --- | --- | --- |
| degrees of freedom | 164 – 13 – 1 = 150 |  | 165 –13– 1 = 151 |  | 183 – 13 – 1 = 169 |  | 185 – 13 – 1 = 171 |  | 184 – 13 – 1 = 170 |  | 149 – 12 – 1 = 136 |  |  |  |
| model mean r | 0.14 |  | 0.14 |  | 0.16 |  | 0.16 |  | 0.17 |  | 0.15 |  |  |  |
| model t values range | -3.09 | 4.78 | -3.42 | 5.83 | -11.12 | 4.05 | -8.33 | 2.93 | -9.69 | 3.12 | -6.31 | 2.63 |  |  |
| t significant treshold after RFT correction | 3.15 |  | 3.15 |  | 3.14 |  | 3.14 |  | 3.14 |  | 3.15 |  |  |  |
| Brain hemisphere | Left | Right | Left | Right | Left | Right | Left | Right | Left | Right | Left | Right | Left | Right |
| amygdala | 0% | 0% | 0% | 0% | 43% | 57% | 14% | 55% | 48% | 73% | 0% | 0% | 1 578 | 1 585 |
| caudal anterior cingulate | 43% | 42% | 87% | 66% | 100% | 100% | 100% | 100% | 100% | 100% | 11% | 8% | 1 641 | 1 383 |
| caudal middle frontal | 45% | 0% | 93% | 76% | 100% | 94% | 100% | 100% | 100% | 100% | 49% | 57% | 4 824 | 4 252 |
| caudate | 3% | 52% | 7% | 53% | 85% | 71% | 40% | 78% | 63% | 76% | 0% | 0% | 2 197 | 2 183 |
| cuneus | 7% | 0% | 27% | 0% | 57% | 29% | 93% | 56% | 93% | 48% | 94% | 61% | 2 539 | 2 644 |
| entorhinal | 2% | 0% | 29% | 1% | 32% | 31% | 4% | 41% | 21% | 43% | 0% | 0% | 1 181 | 986 |
| fusiform | 71% | 0% | 96% | 16% | 100% | 80% | 94% | 97% | 100% | 98% | 14% | 6% | 6 075 | 6 041 |
| hippocampus | 8% | 0% | 36% | 18% | 28% | 51% | 8% | 49% | 26% | 66% | 1% | 0% | 1 915 | 2 050 |
| inferior parietal | 29% | 0% | 96% | 12% | 100% | 92% | 100% | 95% | 100% | 94% | 81% | 67% | 7 608 | 9 594 |
| inferior temporal | 49% | 0% | 87% | 31% | 100% | 96% | 95% | 100% | 100% | 100% | 13% | 0% | 9 568 | 10 355 |
| insula | 65% | 7% | 100% | 81% | 100% | 100% | 99% | 100% | 100% | 100% | 20% | 5% | 5 264 | 5 248 |
| isthmus cingulate | 33% | 0% | 73% | 64% | 100% | 100% | 98% | 100% | 100% | 100% | 97% | 96% | 1 422 | 1 307 |
| lateral occipital | 2% | 0% | 27% | 0% | 70% | 5% | 74% | 18% | 79% | 17% | 64% | 4% | 8 638 | 8 611 |
| lateral orbitofrontal | 78% | 30% | 99% | 93% | 99% | 99% | 99% | 99% | 99% | 99% | 18% | 11% | 3 938 | 4 007 |
| lingual | 13% | 0% | 46% | 6% | 76% | 20% | 84% | 69% | 91% | 46% | 92% | 67% | 3 960 | 4 089 |
| medial orbitofrontal | 83% | 86% | 100% | 99% | 100% | 99% | 100% | 99% | 100% | 99% | 0% | 0% | 2 272 | 2 637 |
| middle temporal | 25% | 0% | 83% | 28% | 100% | 100% | 100% | 100% | 100% | 100% | 55% | 39% | 10 148 | 11 528 |
| pallidum | 0% | 0% | 8% | 0% | 8% | 0% | 0% | 2% | 2% | 0% | 0% | 0% | 1 235 | 1 259 |
| paracentral | 32% | 5% | 64% | 13% | 79% | 55% | 71% | 57% | 73% | 55% | 29% | 20% | 2 564 | 2 248 |
| parahippocampal | 58% | 0% | 82% | 47% | 91% | 87% | 43% | 83% | 82% | 88% | 12% | 3% | 1 292 | 1 213 |
| pars opercularis | 67% | 18% | 93% | 67% | 100% | 100% | 100% | 100% | 100% | 100% | 66% | 77% | 2 357 | 3 140 |
| pars orbitalis | 100% | 32% | 100% | 98% | 100% | 100% | 100% | 100% | 100% | 100% | 52% | 30% | 730 | 605 |
| pars triangularis | 82% | 5% | 99% | 78% | 100% | 100% | 100% | 100% | 100% | 100% | 74% | 69% | 2 828 | 2 723 |
| posterior cingulate | 67% | 23% | 96% | 70% | 100% | 100% | 100% | 100% | 100% | 100% | 89% | 70% | 1 817 | 1 989 |
| pericalcarine | 6% | 0% | 12% | 0% | 66% | 9% | 94% | 21% | 98% | 22% | 100% | 43% | 1 044 | 1 416 |
| postcentral | 13% | 7% | 62% | 45% | 98% | 78% | 92% | 80% | 91% | 74% | 26% | 58% | 5 135 | 4 389 |
| precentral | 24% | 3% | 73% | 49% | 87% | 68% | 83% | 81% | 86% | 79% | 19% | 28% | 6 885 | 6 156 |
| precuneus | 53% | 0% | 91% | 20% | 100% | 95% | 100% | 99% | 100% | 99% | 74% | 81% | 6 680 | 6 715 |
| putamen | 15% | 3% | 80% | 22% | 96% | 62% | 95% | 93% | 97% | 87% | 0% | 0% | 4 930 | 4 915 |
| rostral anterior cingulate | 87% | 95% | 100% | 100% | 100% | 100% | 100% | 100% | 100% | 100% | 0% | 0% | 3 142 | 1 710 |
| rostral middle frontal | 38% | 0% | 91% | 79% | 100% | 100% | 100% | 100% | 100% | 100% | 90% | 98% | 8 243 | 8 210 |
| superior frontal | 50% | 23% | 92% | 51% | 100% | 96% | 100% | 100% | 100% | 99% | 26% | 27% | 14 850 | 17 709 |
| superior parietal | 15% | 0% | 99% | 0% | 100% | 42% | 100% | 85% | 100% | 81% | 10% | 3% | 6 315 | 5 636 |
| supramarginal | 16% | 8% | 94% | 58% | 100% | 93% | 100% | 96% | 100% | 96% | 54% | 79% | 6 265 | 5 221 |
| superior temporal | 53% | 0% | 87% | 56% | 100% | 99% | 92% | 100% | 98% | 100% | 47% | 34% | 8 883 | 8 137 |
| thalamus | 0% | 0% | 11% | 0% | 41% | 0% | 12% | 0% | 34% | 0% | 0% | 0% | 5 305 | 5 261 |
| transverse temporal | 31% | 0% | 100% | 10% | 100% | 100% | 100% | 100% | 100% | 100% | 100% | 91% | 472 | 360 |
| Hemispheric load | 37% | 8% | 77% | 39% | 91% | 78% | 88% | 85% | 92% | 84% | 40% | 34% | 165 740 | 167 512 |
| Significant voxels overlapping DKT atlas | 22% |  | 58% |  | 84% |  | 87% |  | 88% |  | 37% |  | 333 252 |  |
| Total significant voxels in the regression model | 19% |  | 48% |  | 62% |  | 72% |  | 65% |  | 32% |  | 639 106 |  |

**Notes.** Overlap in terms of voxel number between (a) DKT atlas voxels; and (b) Masks of significant association between language scores and tau-PET, in all individuals across the tau continuum, controlling for age + sex + education + APOE genotype + primary language + handedness + mean full cerebellar grey <sup>18</sup>F-AZD-4694 Standardized Uptake Value Ratio i.e. Aβ-PET load; after T-statistical parametric maps were corrected for multiple comparisons using a random field theory cluster threshold of *p* < .001. Maximal possible DKT Atlas (totalling 333 252 voxels) was based on DKT atlas reference substructures voxel count. Maximal total significant voxels (639 106 voxels) was based on mask file upon which the regression is drawn, that is a mask of grey matter within the Whole cortex and Basal ganglia. Cells are colored in accordance with the percentage of significant voxels in a specific region, higher percentages cells have their background colored in a darker shade of green as they tend towards 100%, starting from full white at 0%. DKT: Desikan–Killiany–Tourville, RFT: random field theory.

CU sample-based results are similarly presented in figure 3 and table 4. Association between language scores and tau-PET was significant almost exclusively for complaints over forgetting the names of objects (ECog 1; 17 066 voxels) and over finding the right words to use in a conversation (ECog 3; 12 763 voxels) and to a much lesser extent for confrontation naming (672 voxels). Significant associations were in this case exclusive to the left hemisphere and overlap was concentrated primarily in the inferior temporal and fusiform substructures, but also extended to the parahippocampal, middle temporal, superior temporal, entorhinal, lateral occipital and lingual substructures.

**Figure 3:**
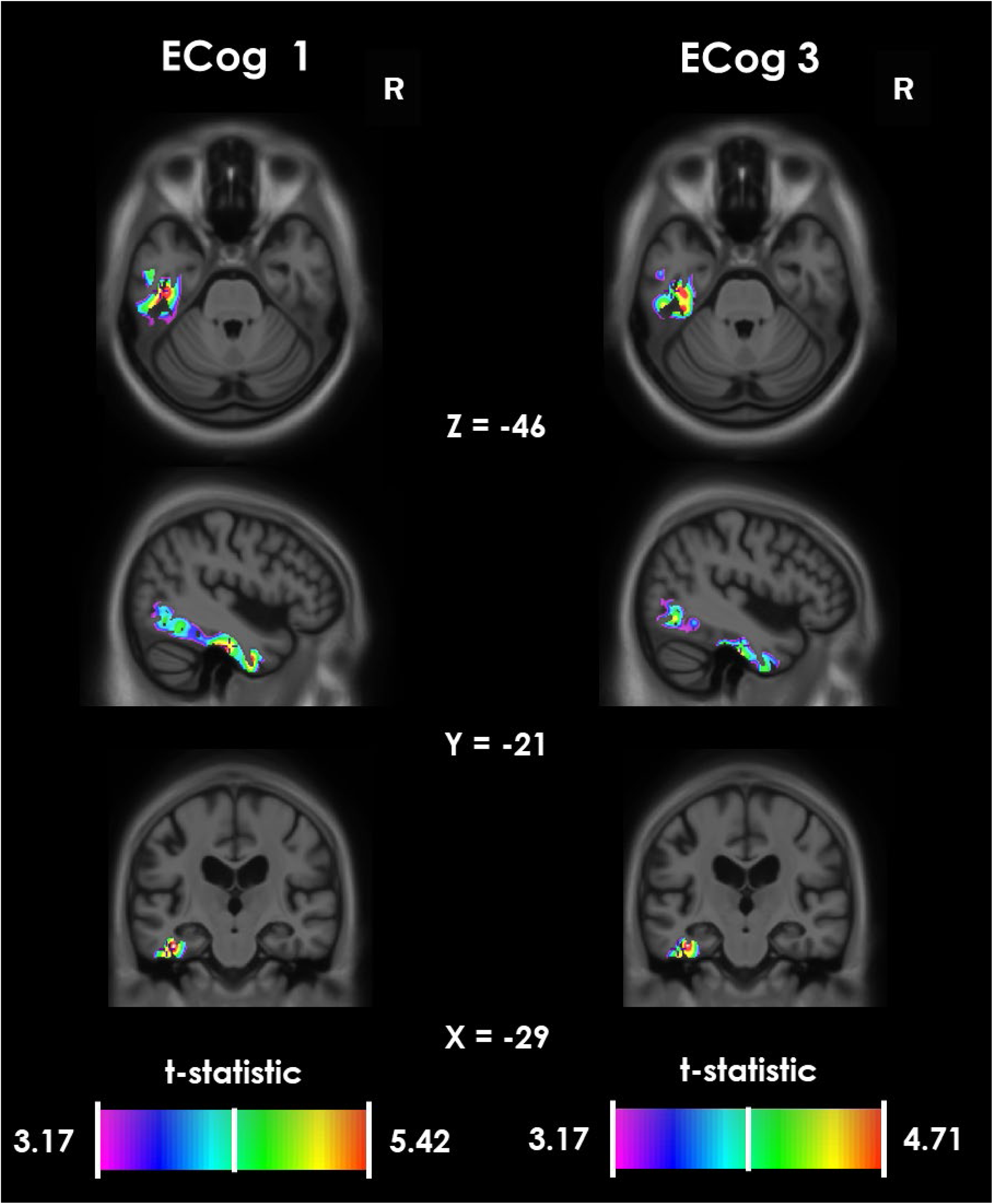
CU sample - Location of significant associations between language scores and tau-PET

**Table 4:** CU sample – Regions of significant associations between language scores and tau-PET

|  | ECog language 1 |  | ECog language 3 |  | Boston Naming Test |  | Phonemic Fluency |  | Semantic Fluency |  | WASI-II Vocabulary |  | Maximal possible<br>DKT Atlas Overlap<br>in number of voxels |  |
| --- | --- | --- | --- | --- | --- | --- | --- | --- | --- | --- | --- | --- | --- | --- |
| degrees of freedom | 109 – 9 – 1 = 99 |  | 110 – 9 – 1 = 100 |  | 125 – 9 – 1 = 115 |  | 126 – 9 – 1 = 116 |  | 125 – 9 – 1 = 115 |  | 107 – 9 – 1 = 97 |  |  |  |
| model mean r | 0.11 |  | 0.11 |  | 0.11 |  | 0.11 |  | 0.11 |  | 0.12 |  |  |  |
| model t values range | -2.58 | 5.42 | -2.76 | 4.71 | -4.11 | 2.57 | -2.30 | 3.44 | -1.85 | 3.95 | -2.52 | 2.22 |  |  |
| t significant treshold after RFT correction | 3.17 |  | 3.17 |  | 3.16 |  | 3.16 |  | 3.16 |  | 3.18 |  |  |  |
| Brain hemisphere | Left | Right | Left | Right | Left | Right | Left | Right | Left | Right | Left | Right | Left | Right |
| amygdala | 0% | 0% | 0% | 0% | 0% | 0% | 0% | 0% | 0% | 0% | 0% | 0% | 1 578 | 1 585 |
| caudal anterior cingulate | 0% | 0% | 0% | 0% | 0% | 0% | 0% | 0% | 0% | 0% | 0% | 0% | 1 641 | 1 383 |
| caudal middle frontal | 0% | 0% | 0% | 0% | 0% | 0% | 0% | 0% | 0% | 0% | 0% | 0% | 4 824 | 4 252 |
| caudate | 0% | 0% | 0% | 0% | 0% | 0% | 0% | 0% | 0% | 0% | 0% | 0% | 2 197 | 2 183 |
| cuneus | 0% | 0% | 0% | 0% | 0% | 0% | 0% | 0% | 0% | 0% | 0% | 0% | 2 539 | 2 644 |
| entorhinal | 5% | 0% | 7% | 0% | 0% | 0% | 0% | 0% | 0% | 0% | 0% | 0% | 1 181 | 986 |
| fusiform | 71% | 0% | 38% | 0% | 0% | 0% | 0% | 0% | 0% | 0% | 0% | 0% | 6 075 | 6 041 |
| hippocampus | 0% | 0% | 0% | 0% | 0% | 0% | 0% | 0% | 0% | 0% | 0% | 0% | 1 915 | 2 050 |
| inferior parietal | 0% | 0% | 0% | 0% | 0% | 0% | 0% | 0% | 0% | 0% | 0% | 0% | 7 608 | 9 594 |
| inferior temporal | 63% | 0% | 53% | 0% | 0% | 0% | 0% | 0% | 0% | 0% | 0% | 0% | 9 568 | 10 355 |
| insula | 0% | 0% | 0% | 0% | 0% | 0% | 0% | 0% | 0% | 0% | 0% | 0% | 5 264 | 5 248 |
| isthmus cingulate | 0% | 0% | 0% | 0% | 0% | 0% | 0% | 0% | 0% | 0% | 0% | 0% | 1 422 | 1 307 |
| lateral occipital | 3% | 0% | 5% | 0% | 0% | 0% | 0% | 0% | 0% | 0% | 0% | 0% | 8 638 | 8 611 |
| lateral orbitofrontal | 0% | 0% | 0% | 0% | 0% | 0% | 0% | 0% | 0% | 0% | 0% | 0% | 3 938 | 4 007 |
| lingual | 5% | 0% | 0% | 0% | 0% | 0% | 0% | 0% | 0% | 0% | 0% | 0% | 3 960 | 4 089 |
| medial orbitofrontal | 0% | 0% | 0% | 0% | 0% | 0% | 0% | 0% | 0% | 0% | 0% | 0% | 2 272 | 2 637 |
| middle temporal | 8% | 0% | 5% | 0% | 0% | 0% | 0% | 0% | 0% | 0% | 0% | 0% | 10 148 | 11 528 |
| pallidum | 0% | 0% | 0% | 0% | 0% | 0% | 0% | 0% | 0% | 0% | 0% | 0% | 1 235 | 1 259 |
| paracentral | 0% | 0% | 0% | 0% | 0% | 0% | 0% | 0% | 0% | 0% | 0% | 0% | 2 564 | 2 248 |
| parahippocampal | 14% | 0% | 8% | 0% | 0% | 0% | 0% | 0% | 0% | 0% | 0% | 0% | 1 292 | 1 213 |
| pars opercularis | 0% | 0% | 0% | 0% | 0% | 0% | 0% | 0% | 0% | 0% | 0% | 0% | 2 357 | 3 140 |
| pars orbitalis | 0% | 0% | 0% | 0% | 0% | 0% | 0% | 0% | 0% | 0% | 0% | 0% | 730 | 605 |
| pars triangularis | 0% | 0% | 0% | 0% | 0% | 0% | 0% | 0% | 0% | 0% | 0% | 0% | 2 828 | 2 723 |
| posterior cingulate | 0% | 0% | 0% | 0% | 0% | 0% | 0% | 0% | 0% | 0% | 0% | 0% | 1 817 | 1 989 |
| pericalcarine | 0% | 0% | 0% | 0% | 0% | 0% | 0% | 0% | 0% | 0% | 0% | 0% | 1 044 | 1 416 |
| postcentral | 0% | 0% | 0% | 0% | 0% | 0% | 0% | 0% | 0% | 0% | 0% | 0% | 5 135 | 4 389 |
| precentral | 0% | 0% | 0% | 0% | 0% | 0% | 0% | 0% | 0% | 0% | 0% | 0% | 6 885 | 6 156 |
| precuneus | 0% | 0% | 0% | 0% | 0% | 0% | 0% | 0% | 0% | 0% | 0% | 0% | 6 680 | 6 715 |
| putamen | 0% | 0% | 0% | 0% | 0% | 0% | 0% | 0% | 0% | 0% | 0% | 0% | 4 930 | 4 915 |
| rostral anterior cingulate | 0% | 0% | 0% | 0% | 0% | 0% | 0% | 0% | 0% | 0% | 0% | 0% | 3 142 | 1 710 |
| rostral middle frontal | 0% | 0% | 0% | 0% | 0% | 0% | 0% | 0% | 0% | 0% | 0% | 0% | 8 243 | 8 210 |
| superior frontal | 0% | 0% | 0% | 0% | 0% | 0% | 0% | 0% | 0% | 0% | 0% | 0% | 14 850 | 17 709 |
| superior parietal | 0% | 0% | 0% | 0% | 0% | 0% | 0% | 0% | 0% | 0% | 0% | 0% | 6 315 | 5 636 |
| supramarginal | 0% | 0% | 0% | 0% | 0% | 0% | 0% | 0% | 0% | 0% | 0% | 0% | 6 265 | 5 221 |
| superior temporal | 6% | 0% | 8% | 0% | 0% | 0% | 0% | 0% | 0% | 0% | 0% | 0% | 8 883 | 8 137 |
| thalamus | 0% | 0% | 0% | 0% | 0% | 0% | 0% | 0% | 0% | 0% | 0% | 0% | 5 305 | 5 261 |
| transverse temporal | 0% | 0% | 0% | 0% | 0% | 0% | 0% | 0% | 0% | 0% | 0% | 0% | 472 | 360 |
| Hemispheric load | 7% | 0% | 6% | 0% | 0% | 0% | 0% | 0% | 0% | 0% | 0% | 0% | 165 740 | 167 512 |
| Significant voxels overlapping DKT atlas | 4% |  | 3% |  | 0% |  | 0% |  | 0% |  | 0% |  | 333 252 |  |
| Total significant voxels in the regression model | 3% |  | 2% |  | 672 (0%) |  | 0% |  | 0% |  | 0% |  | 639 106 |  |

**Notes.** Overlap in terms of voxel number between (a) DKT atlas voxels; and (b) Masks of significant association between language scores and tau-PET, in cognitively unimpaired individuals, controlling for age + sex + education + APOE genotype + primary language + handedness + mean full cerebellar grey <sup>18</sup>F-AZD-4694 Standardized Uptake Value Ratio i.e. Aβ-PET load; after T-statistical parametric maps were corrected for multiple comparisons using a random field theory cluster threshold of *p* < .001. Maximal possible DKT Atlas (totalling 333 252 voxels) was based on DKT atlas reference substructures voxel count. Maximal total significant voxels (639 106 voxels) was based on mask file upon which the regression is drawn, that is a mask of grey matter within the Whole cortex and Basal ganglia. Cells are colored in accordance with the percentage of significant voxels in a specific region, higher percentages cells have their background colored in a darker shade of green as they tend towards 100%, starting from full white at 0%. DKT: Desikan–Killiany–Tourville, RFT: random field theory.

### 3.3 Prediction of early AD amyloid and tau biological stage

Models comparing the importance of previously used language variables and covariates, with the inclusion of the MoCA screening tool, in the classification of participants into amyloid and tau based early AD pathology groups (Braak 0 Aβ–, Braak 0 Aβ+, Braak 1-2 Aβ+) are presented in table 5 In order of importance, complaints over forgetting the names of objects (ECog 1), APOE genotype and confrontation naming significantly contributed to the classification of participants into their respective early AD subgroup.

**Table 5:** Best predictors of early AD pathology

| Classification for : | Braak 0 [Aβ negative]<br>Braak 0 [Aβ positive]<br>Braak 1-2 [Aβ positive] | Singular Pillai | Pillai advancement<br>Threshold |
| --- | --- | --- | --- |
| **Model 1 (with ECog language 1)** | | Overall Pillai's trace = .32, $p < .001$ | |
|  | Ecog language 1: Forgetting the names of objects* | .16 | .07 |
|  | APOE genotype* | .09 | .07 |
|  | Boston Naming Test total score* | .07 | .07 |
|  | **MoCA total score** | **.04** |  |

**Notes.** \*: Significant to the prediction model. Alpha threshold for Pillai's advancement was set to $p = .05$ . Analysis of a model without alpha threshold indicated that MoCA followed BNT in all models, but did not meet the threshold for variable selection. This was followed by Sex; Ecog 3; Education; Semantic Fluency; Age; WASI-II; Handedness; Primary Language and Phonemic Fluency.

## 4. Discussion

The present study aimed to bridge the clinical and biological AD continuum definitions by utilizing language performance data from a well-characterized sample of participants at different stages of pathological tau accumulation according to a PET-based Braak staging framework. Our observations highlight a progression of language impairments which parallels tau’s progressive accumulation within the brain. The earliest stages of tau progression (Braak 1-2) were characterized by elevated complaints over forgetting the names of objects, followed by objective impairments in confrontation naming in the middle stages (Braak 3-4) and further widespread impairments of semantic fluency, phonemic fluency, word-knowledge processes as well elevated complaints over finding the right words to use in a conversation in the final stages (Braak 5-6). When investigating composite language, screening, demographic and genetic measures, complaints over forgetting the names of objects and impairments in confrontation naming were once again, along with APOE genotype, the only variables significantly contributing to the correct classification of participants into groups of early AD pathology (Braak 0 Aβ–, Braak 0 Aβ+ and Braak 1-2 Aβ+). Across the whole AD continuum, imaging results then highlighted widespread associations between tau aggregation and all language measures. Even more importantly, in the cognitively unimpaired, associations were nearly exclusive to word-finding complaints and centered around the left fusiform and inferior temporal gyri. Overall, these findings permit the staging of important language symptoms across the core biological definition of AD and identify relevant clinical measures for the cost-effective detection of early pathology and tracking of disease progression.

Consistent with hypothesis 1 and previously highlighted language studies primarily based around clinically defined AD, we observed elevated word-finding complaints as well as impairments of confrontation naming, phonemic fluency, semantic fluency and word-knowledge processes along the biologically defined AD continuum. Similar to studies which investigated subjective memory complaints and found it was sensitive to earlier decline when compared to objective measurements (Mitchell et al., 2014; Pike et al., 2022), we found that complaints over forgetting the names of objects were more sensitive to early AD pathology when compared to objective language measures (confrontation naming, fluencies, word-knowledge). Also consistent with previous findings, confrontation naming impairments appeared from the middle stages of tau progression (Braak 3-4), at the same time that biologically defined subgroups within our sample became predominantly MCI. This was despite picture naming tests, and specifically the BNT, not being the most ideal or sensitive measure of AD-related lexicosemantic deficits (Harry and Crowe, 2014; Joubert et al., 2021) which suggests that better measures, such as naming tasks involving unique exemplars like that of famous people, historical events, places or buildings (Montembeault et al., 2017; Joubert et al., 2021) could be even better suited at detecting early pathology or at tracking between-stages progression of tau along its continuum. In contrast with previous studies (Nutter-Upham et al., 2008; Chasles et al., 2020) neither phonemic nor semantic fluencies were found to be impaired from the middle stages of tau progression and instead, most of the overall loss in word production for each measure was only observed within the last stages (Braak 5-6). This could be due to the many processes involved (besides language) in verbal fluency tasks such as executive function, processing speed and perception (Joubert et al., 2021). Fluency may therefore suffer from our controlling of covariates of age, sex and education in univariate models (Nogueira et al., 2016). In semantic fluency, divergence in results could additionally be explained by task differences, as total scores for our sample included animals’ names similar to previous works, with the addition of boys’ names which was more unusual. This is particularly relevant considering concerns in the literature that fluency performance could be condition-dependant, with varying results based on the categories or letters used for the task (Rinehardt et al., 2014). Also in line with previous findings, word-knowledge was not preserved in the final stages of tau progression (Braak 5-6; Gontkovsky et al., 2022). This test involves AD-sensitive semantic performance (Marier et al., 2024) however, it also allows for circumlocutions (which confrontation naming attempts to bypass), it’s low sensitivity to early pathology therefore comes to no surprise. Consistent with our second hypothesis, our multivariate results captured the impacts of demographic measures across the biologically defined AD continuum. Most importantly, APOE genotype, perhaps better captured through our ordinal computation, was the variable most linked to a model proportional to tau’s progression, consistent with APOE’s established impact on the risk of developing late-onset AD (Qian et al., 2017; Reiman et al., 2020; Sienski et al., 2021). Additionally, and consistent with our fourth hypothesis, alongside APOE genotype, complaints over forgetting the names of objects and confrontation naming were the only language variables significantly contributing to the correct classification of cognitively unimpaired participants into groups of early AD pathology (Braak 0 Aβ–, Braak 0 Aβ+ and Braak 1-2 Aβ+), being firstmost and thirdmost contributing respectively. Taken together, these results underscore the need for further investigation into language measures involving word-finding complaints and confrontation naming. These measures have shown that they can be useful in the early detection of AD pathology and can track to specific stages of its development, despite their severely limited number of items (particularly in the case of word-finding complaints) and in spite also of the lag between test development and implementation into clinical practice.

Consistent with our third hypothesis and echoing previous results linking language measures to AD-related GM atrophy, we found associations (a) between words-finding complaints and the left fusiform gyrus (Montembeault et al., 2022). However, our results extended to both complaints measures and extended further in the inferior temporal, parahippocampal and superior temporal gyri in CU and even beyond in pars orbitalis and temporal regions when considering the whole tau continuum and were particularly widespread for complaints over finding the right words to use in a conversation; (b) between confrontation naming, the left anterior temporal lobe (Frings et al., 2011; Domoto-Reilly et al., 2012) and left inferior prefrontal gyrus (Joubert et al., 2010). However, our results also extended bilaterally to frontal, temporal and parietal regions, with other important associations (aside from left temporal) in the left orbitofrontal cortex and left superior frontal gyrus; (c) between verbal fluency, the left inferior temporal gyrus, left insular cortex, bilateral hippocampus and parahippocampal gyrus, as well as with the anterior and posterior cingulate cortices and bilateral caudate nucleus (Rodríguez-Aranda et al., 2016). However, our results were much more widespread than previously found and extended to frontal and parietal regions. This was particularly the case for phonemic fluency for which relationships with tau extended to virtually the same regions as semantic fluency. Differences between fluencies included more specific involvement of the left entorhinal cortex, hippocampus, parahippocampal gyrus, amygdala and thalamus in semantic fluency and right lingual gyrus in phonemic fluency. We also found significant associations between tau and word-knowledge processes, centered around occipital, temporal and frontal regions, particularly in the left isthmus cingulate and bilateral rostral middle frontal, which to the best of our knowledge had not previously been documented in older adults. Overall these results are consistent with classical and recent neuroanatomical models of a left-weighted language network spanning the anterior temporal lobe, inferior frontal gyrus, posterior middle temporal gyrus and temporo-parietal junction, with fronto-temporal, parieto-temporal, occipito-temporal and fronto-frontal connections (Geschwind, 1970; Tremblay and Dick, 2016). Widespread relationships are also in line with the many processes involved besides language, particularly in confrontation naming (Harry and Crowe, 2014) and fluencies (Joubert et al., 2021). These results are also consistent with previous findings which highlighted relationships between language performance measures and AD-related GM atrophy as well as between word-finding complaints and increased levels of Aβ, with these relationships now being extended to tau protein aggregation within the brain.

While the current study fulfills many gaps in the literature, these results also need to be considered within the context of several limitations. First, the cross-sectional design of the study does not confirm that word-finding complaints and objective language impairments evolve with time across the biologically defined AD continuum, as a longitudinal design would. Second, combining Braak stages may have affected results dependent on group comparisons. Third, important risk factors could not be accounted for despite their reported relevance such as those related social, physical and cognitive activity both in the mid and later life (Livingston et al., 2024). Fourth, the biological AD continuum is not limited to Aβ and tau detected using ^18^F-MK-6240 and ^18^F-AZD-4694 radioligand in PET, nor is it limited to the brain’s grey matter (Jack et al., 2024) and additionally, most cases of dementia involve mixed pathologies (Alzheimer’s disease facts and figures, 2025), this implies that area-specific binding and disregard of mixed pathologies may have affected our results and limited their clinical utility. Sixth, whilst most of the limitations of the language measures used within this study have been discussed, other important domains of language have been neglected, such a those involving connected speech features (Slegers et al., 2018). Further research is therefore essential to expand upon our findings.

In conclusion, our results lend support to the usefulness of subjective word-finding complaints over forgetting the names of objects when assessing the likelihood of underlying Alzheimer’s Disease at the earliest stages of AD pathology. The belief that this ability has been worsening should therefore not be discredited on the sole basis that it involves subjectivity. Similarly to genetic risks (which it outperformed in our early AD classification models), it instead appears to be accompanied with significantly raised risks of underlying AD pathology in accordance with the strength of the perceived impairment. Results also lend credence to calls for more sensitive language measures to be developed, particularly in the case of word-finding complaints, and for already existing more sensitive tests to be implemented within clinical settings.

## Data Availability

All the data collected under the TRIAD cohort is governed by the policies set by the Research Ethics Board Office of the McGill University, Montreal and the Douglas Research Center, Verdun.
If you are a researcher and would like to access the TRIAD data for research purpose, please submit a data share request via the appropriate data share channels (https://triad.tnl-mcgill.com/contact-us/) or seek out the corresponding author if you required help in this process.

## Abbreviations

Aβ: Amyloid beta
AD: Alzheimer’s Disease
ADNI: Alzheimer’s Disease Neuroimaging Initiative
ANCOVA: Analyses of covariance
ANOVA: Analyses of variance
ANTs: Advanced Normalization Tools
BNT: Boston Naming Test
CDR: Clinical Dementia Rating
CU: Cognitively unimpaired
DKT: Desikan–Killiany–Tourville
ECog: Everyday Cognition questionnaire
GM: Grey matter
HEAD: Head-to-head Evaluation of tau tracers in Alzheimer’s Disease
MCI: Mild cognitive impairment
MNI: Montréal Neurological Institute
MoCA: Montréal Cognitive Assessment
MRI: Magnetic resonance imaging
OSEM: Ordered-subsets expectation maximization algorithm
PET: Positron emission tomography
SUVR: Standardized uptake value ratios
TNL: Translational Neuroimaging Laboratory
TRIAD: Translational Biomarkers of Aging and Dementia
WASI-II: Wechsler Abbreviated Scale of Intelligence-II

## Ethics declarations

The TRIAD cohort study was conducted following the ethical standards of all involved institutional research committees and adheres to the principles outlined in the 1975 Helsinki Declaration and its later amendments. The study was approved by the Douglas Mental Health University Institute Research Ethics Board and written informed consent was obtained from all participants or their caregivers.

## Competing interests

AM, JFA, EA, BJH, ACM, NR, GB declare that they have no competing interests relevant to the context of this manuscript.

PV has been affiliated with NovoNordisc, Eisai, Lilly and IntelGenx Corp.

PRN has been affiliated with Roche, Cerveau Radiopharmaceuticals, Lilly, Eisai, Pfizer, Novo Nordisk, and Biogen.

## Contributions

AM: Conceptualization (support); Data Curation (lead); Formal Analysis (lead); Methodology (lead); Project Administration (lead); Software (support); Validation (lead); Visualization (lead); Writing – original draft preparation (lead); Writing – review & editing (support).

JFA: Conceptualization (support); Methodology (support); Resources (support); Software (support); Supervision (support).

EA: Formal Analysis (support); Methodology (support); Resources (support); Software (support); Supervision (support); Validation (support).

BJH: Formal Analysis (support); Methodology (support); Resources (support); Software (support); Supervision (support).

ACM: Conceptualization (support); Methodology (support); Project Administration (support); Resources (support); Supervision (support); Validation (support).

NR: Conceptualization (support); Methodology (support); Supervision (support).

GB: Formal Analysis (support); Methodology (support); Software (support); Resources (support); Supervision (support).

PV: Conceptualization (support)

PRN: Conceptualization (support); Methodology (lead); Resources (lead); Supervision (lead); Writing – Review & Editing (support).

MM: Conceptualisation (support); Project administration (lead); Funding Acquisition (lead); Methodology (lead); Resources (support); Supervision (lead); Validation (support); Writing – Review & Editing (lead).

