## Supplemental Table 1 and 2 for "Language deficits across PET-based Braak stages of tau accumulation in Alzheimer’s disease"

### Supplementary materials

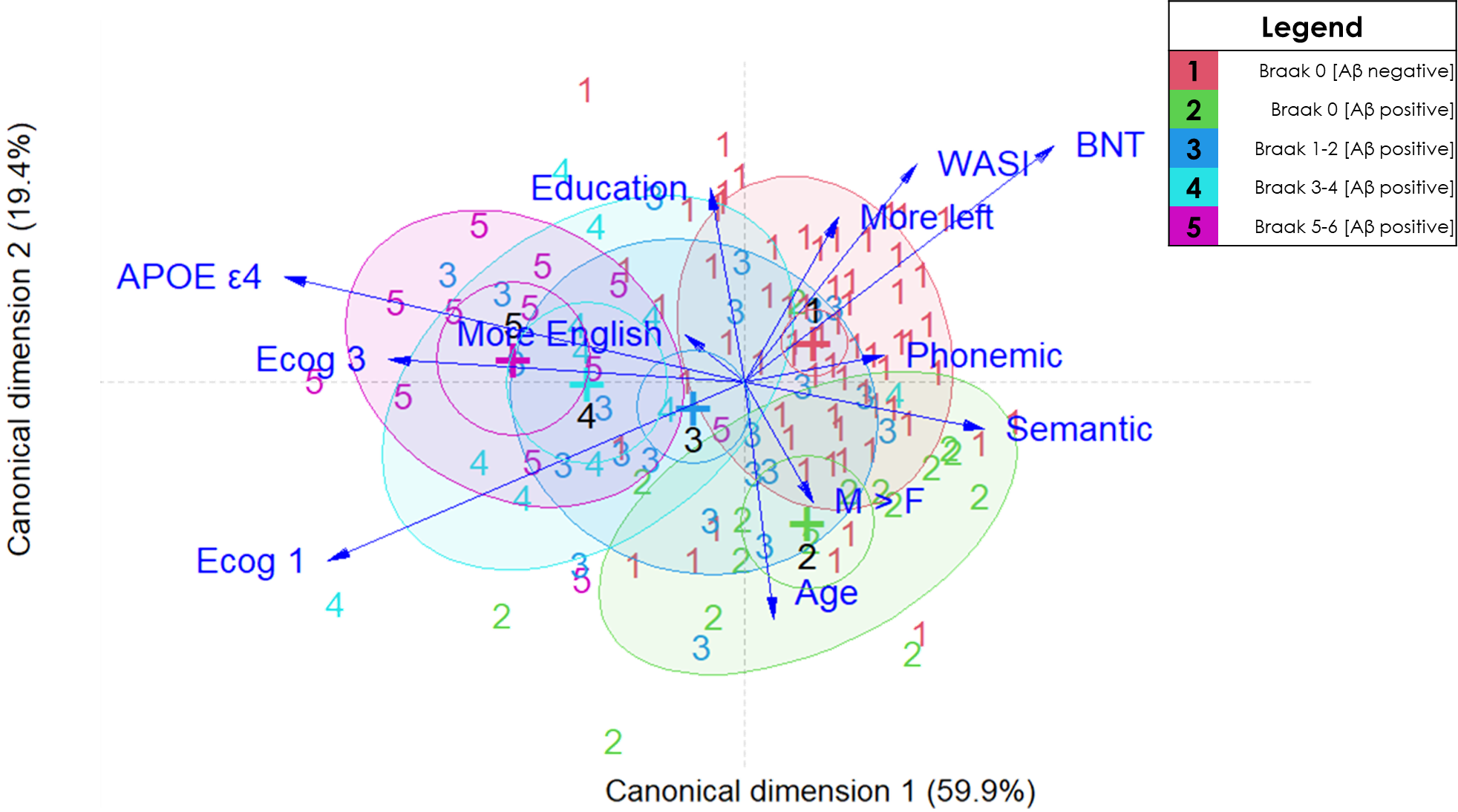

**Supplementary Figure 1**
Visualisation of the direct discriminant analysis results

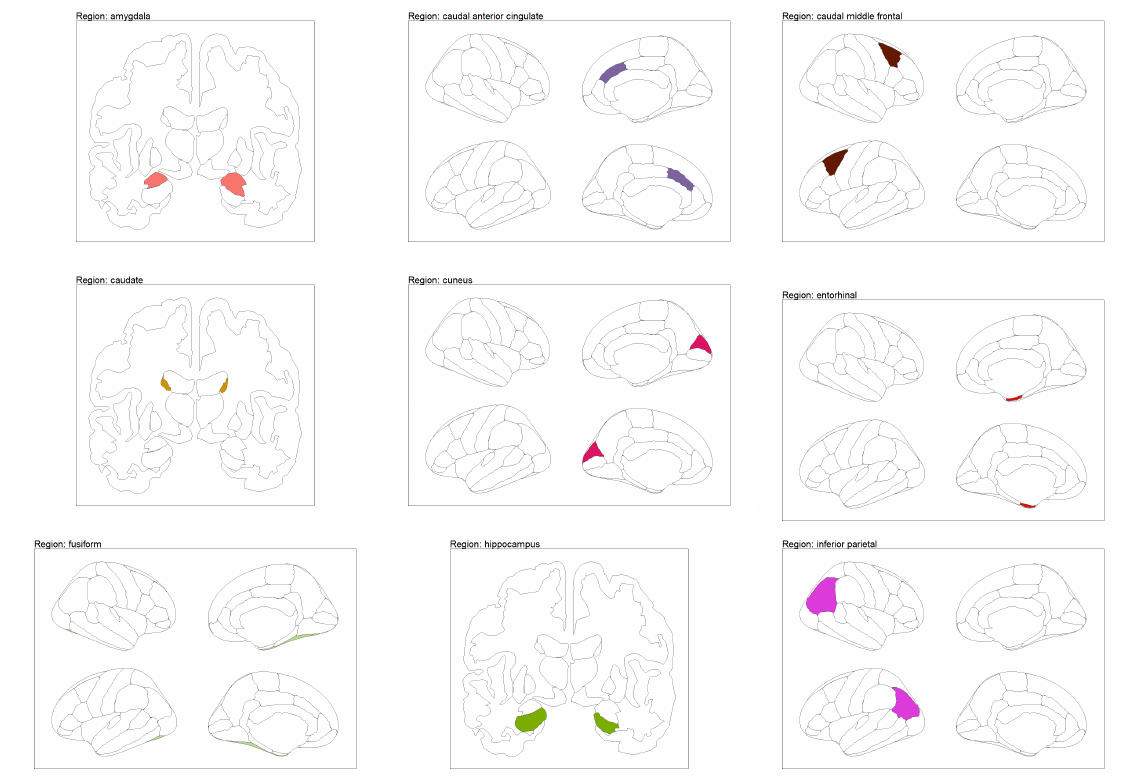

**Supplementary Figure 2**
Reference for DKT atlas brain regions
A. Part 1 of 5

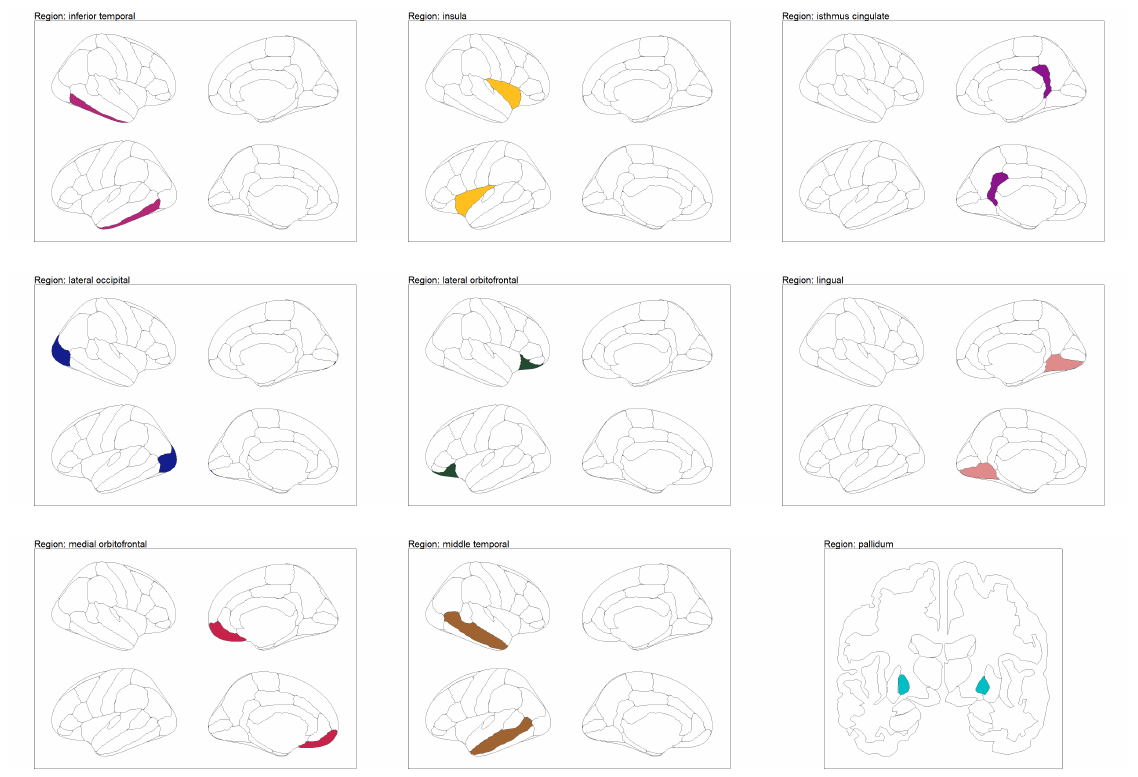

**Supplementary Figure 2**
Reference for DKT atlas brain regions
B. Part 2 of 5

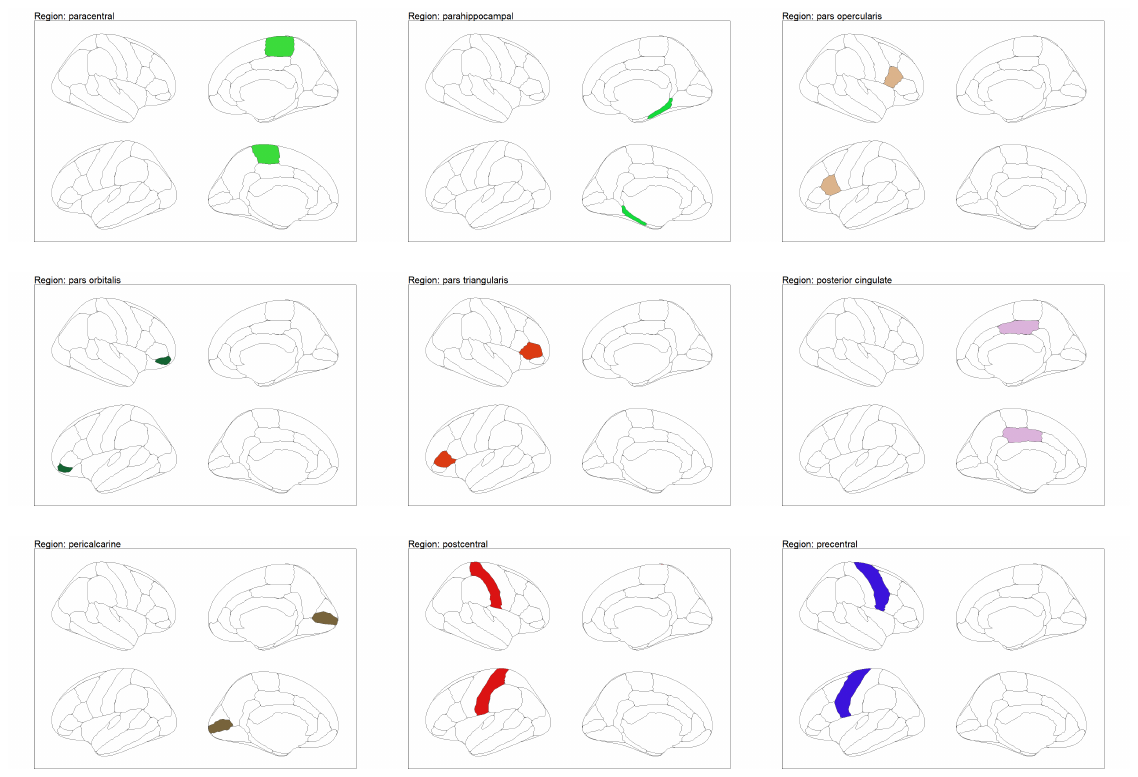

**Supplementary Figure 2**
Reference for DKT atlas brain regions
C. Part 3 of 5

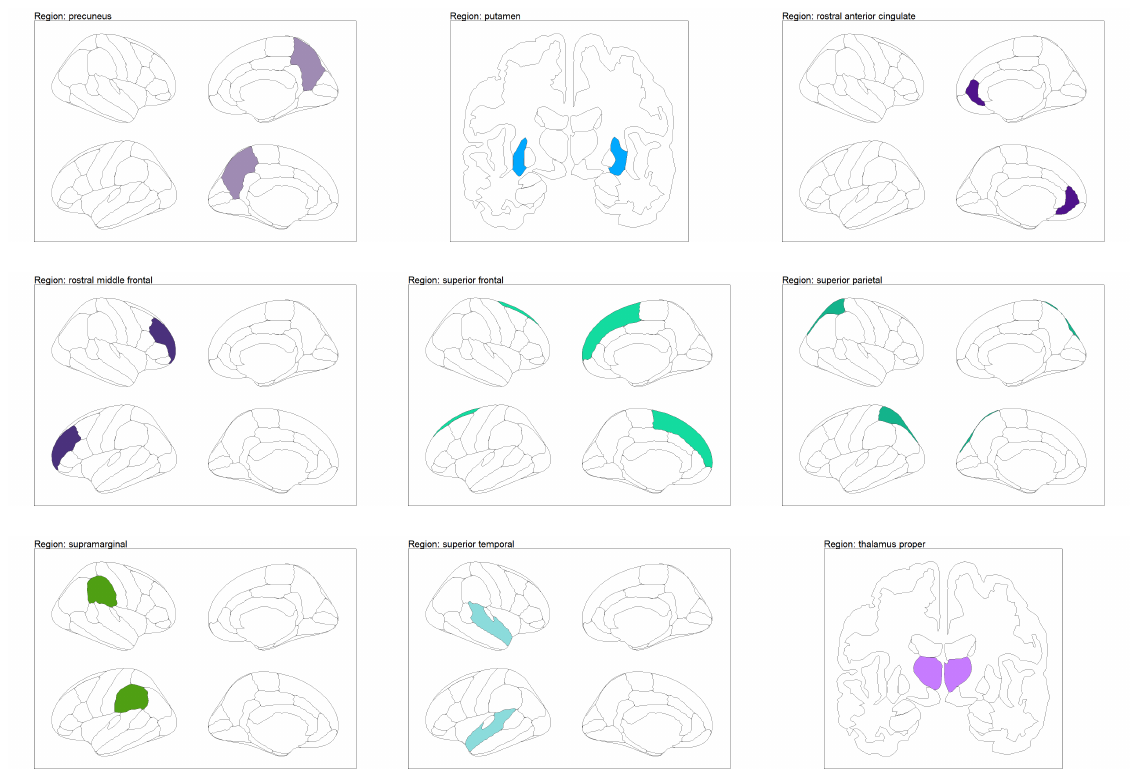

**Supplementary Figure 2**
Reference for DKT atlas brain regions
D. Part 4 of 5

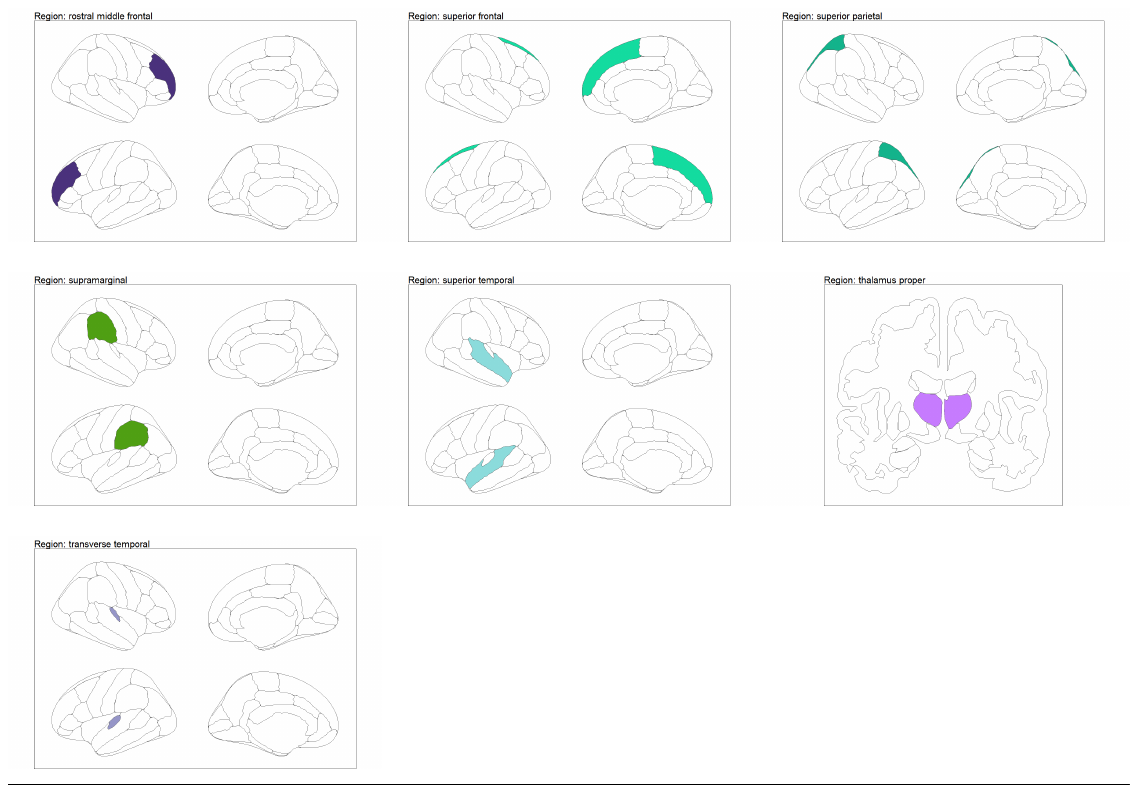

**Supplementary Figure 2**
Reference for DKT atlas brain regions
E. Part 5 of 5
